# Modeling MRSA decolonization: Interactions between body sites and the impact of site-specific clearance

**DOI:** 10.1101/2020.11.30.20239798

**Authors:** Onur Poyraz, Mohamad R. A. Sater, Loren G. Miller, James A. McKinnell, Susan S. Huang, Yonatan H. Grad, Pekka Marttinen

## Abstract

Methicillin-resistant *Staphylococcus aureus* (MRSA) can colonize multiple body sites, and carriage is a risk factor for infection. Successful decolonization protocols reduce disease incidence; however, multiple protocols exist, comprising diverse therapies targeting multiple body sites, and the optimal protocol is unclear. Standard methods are inadequate to infer the impact of site-specific components on successful decolonization. Here, we formulate a Bayesian coupled hidden Markov model (CHMM), which estimates interactions between body sites, quantifies the contribution of each therapy to successful decolonization, and enables predictions of the efficacy of therapy combinations. We applied the model to longitudinal data from a randomized controlled trial (RCT) of an MRSA decolonization protocol consisting of chlorhexidine body and mouthwash and nasal mupirocin. Our findings 1) confirmed nares as a central hub for MRSA colonization and nasal mupirocin as the most crucial therapy, and 2) demonstrated that all components contributed significantly to the efficacy of the protocol and the protocol reduced self-inoculation. Finally, we assessed the impact of hypothetical increases on the effectiveness of each therapy *in silico* and found that enhancing MRSA clearance at the skin would yield the largest gains to the overall decolonization regimen. This study demonstrates the use of advanced modeling to go beyond what is typically achieved by RCTs, enabling evidence-based decision-making to streamline clinical protocols.

## 1 Introduction

Methicillin-resistant *Staphylococcus aureus* (MRSA) is a common antimicrobial-resistant pathogen in community and healthcare settings [1, 2], causing an estimated 320,000 infections in hospitalized patients and over 10,000 deaths in the United States in 2017 [3]. Progress in reducing invasive MRSA infections has slowed, underscoring the importance of continued innovation and effort to prevent disease [4]. As MRSA carriage is a major risk factor for invasive disease, efforts at prevention center on the promotion of decolonization protocols and body hygiene as well as environmental cleaning [5]. The most common *S. aureus* carriage site is the anterior nares, but MRSA can also colonize the perineum and groin, the axilla, the pharynx, as well as other body sites [6, 7]. While the anterior nares have been identified as a key reservoir for transmission and nasal colonization is a major risk factor for invasive disease [8], the extent of interaction among colonization sites and the importance of additional decolonization products targeting other body sites, and the value of increasing their adherence in decolonization protocols remain unclear. It would be ideal to understand the interactions between body sites and the attributable effect of each therapy on overall body clearance. For example, if carriage at a particular body site appears to be dependent on carriage at other body sites, therapies targeting the influencing site would be more efficient, while therapies targeting the dependent site might be less effective for reducing the total body carriage. Achieving this goal requires a detailed understanding of the dynamic relationships of colonization between and among sites.

The CLEAR (Changing Lives by Eradicating Antibiotic Resistance) Trial demonstrated that the use of a post-discharge decolonization protocol in MRSA carriers reduces infection and hospitalization rates [9]. In the trial, 2,121 study participants were randomized into two groups to test the impact of the decolonization protocol: the *education* group (n=1,063) received an educational binder on hygiene, cleanliness, and MRSA transmission; the *decolonization* group (n=1,058) received the same information and as well underwent decolonization protocol for five days twice monthly for six months, with the protocol consisting of nasal mupirocin and chlorhexidine body and mouth wash. During these six months, swabs were collected from the participants at discharge from the hospitalization and three follow-up visits, which approximately took place at months 1, 3, and 6 after the discharge. Samples were taken from the nares, skin (axilla/groin), throat, and, if present, any wound. Participants had different numbers of observations because of trial exits or skipped visits. Overall, 20,506 samples were received in the first 6 months of follow-up. Additionally, the dataset included reported adherence to the protocol, enabling the assessment of both real-world uptake and consideration of the ideal scenario of full compliance.

In this paper, our goal was to model the process of MRSA carriage, with and without the decolonization protocol. Successful modeling can enable characterization of the interactions among MRSA colonization at different body sites and the efficiency of each protocol component. Using this information, we can predict how the decolonization protocol could be more efficient. To achieve these goals, we used a coupled hidden Markov model (CHMM [10, 11, 12, 13, 14, 15, 16], an extension of the standard hidden Markov model, HMM [17, 18, 19]), where the probability of colonization at a particular body site in the next step depends not only on the colonization of the same site but also on the colonization of the other body sites (see Figure 1). We developed a novel formulation of the CHMM: *Additive-CHMM* where the probability of colonization at a particular site is an additive function of colonization at the other sites. According to a predictive model selection criterion (leave-one-out cross-validation, LOO-CV [20]), the Additive-CHMM had superior accuracy compared to a set of site-specific standard HMMs and other formulations that we developed (see Supplementary Table 1). Consequently, the outcomes of the Additive-CHMM are presented and discussed in the main text. Models are described in the Methods and their advantages and disadvantages are discussed in detail in the Supplementary Materials. We provide a practical API as an R-package that implements these models with an efficient Metropolis-within-Gibbs Markov chain Monte Carlo (MCMC) algorithm, which yields Bayesian credible intervals (CI) for all model parameters (see Methods).

**Figure 1.**
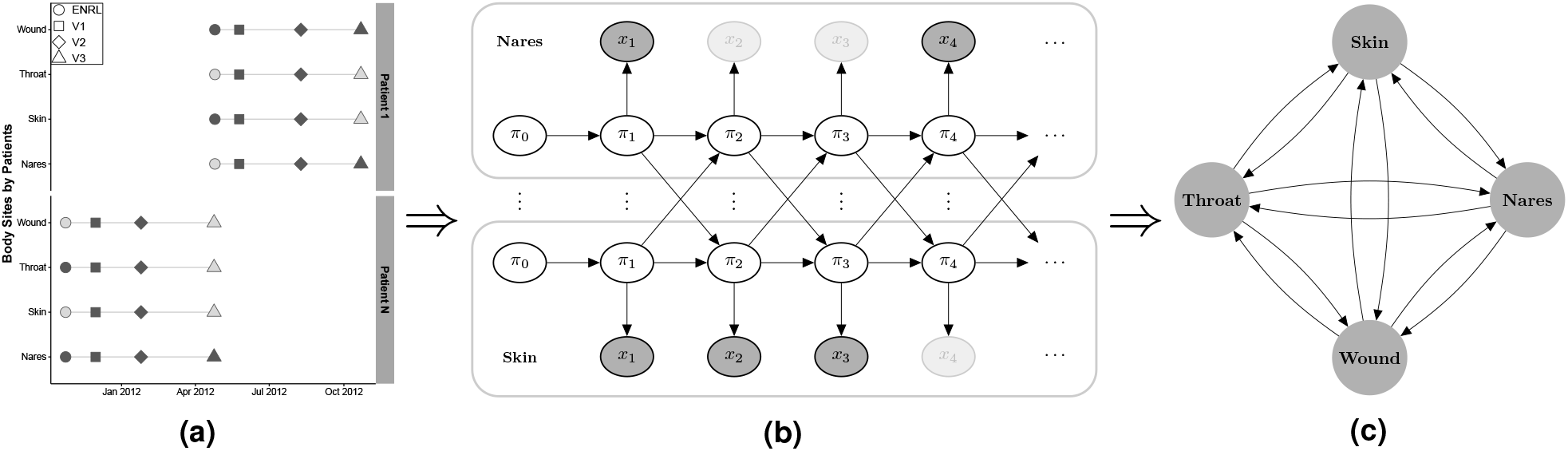
Overview of the modeling strategy. (a) Visit records, (b) illustration of the coupled hidden Markov model (CHMM), and (c) estimated interactions between body sites. **(a)** An example with visit records for two participants. Evaluated visits approximately took place in 1 (V1), 3 (V2), and 6 (V3) months after enrollment (ENRL) in the trial. Filled markers correspond to the collected swabs, and faded markers represent samples missing due to trial exits or skipped visits [9]. **(b)** Here, for clarity, the CHMM is illustrated with two (nares and skin) of the four body sites (nares, skin, throat, and wound). In practice, the model includes all four sites. *π*_*t*_s represent the unobserved true states (whether the site was colonized or not), and *x*_*t*_ represent observed states (whether MRSA was detected from the swab or not) at time *t*. The faded observation nodes correspond to missing observations, which are straightforward to analyze with the CHMM. The sequential model can be used to predict the dynamics of carriage. **(c)** The CHMM allows us to estimate and visualize interaction dynamics graphically, where edges represent the strength and direction of the interaction.

## 2 Results

### The CHMM accurately predicted the decrease in MRSA carriage over the study period

The key metric of success for our model was the extent to which it recapitulates the clearance in MRSA carriage over the study period. To examine this posterior predictive checking, we 1) estimated the parameters of the CHMM in both Education and Decolonization groups, then 2) simulated patient trajectories using parameters from the estimated models, and finally, 3) compared the reduction in colonization in our model-based simulations to the observed changes in the CLEAR trial data. The predicted reduction of the carriage for individuals was from 61% (90% CI is 59%-63%) to 47% (44%-50%) in the education group and down to 25% (22%-28%) in the decolonization group, which accurately matched the observations (see Figure 2). Moreover, the predicted reduction at any single site was accurate as well. To further validate the model’ s ability to capture the dynamics of MRSA carriage, we 1) applied cross-validation (CV) test to inspect the predictive performance (see Supplementary Figure 1), and 2) tested the model using synthetic data to inspect the ability to recover the true parameters (see Supplementary Figure 2).

**Figure 2.**
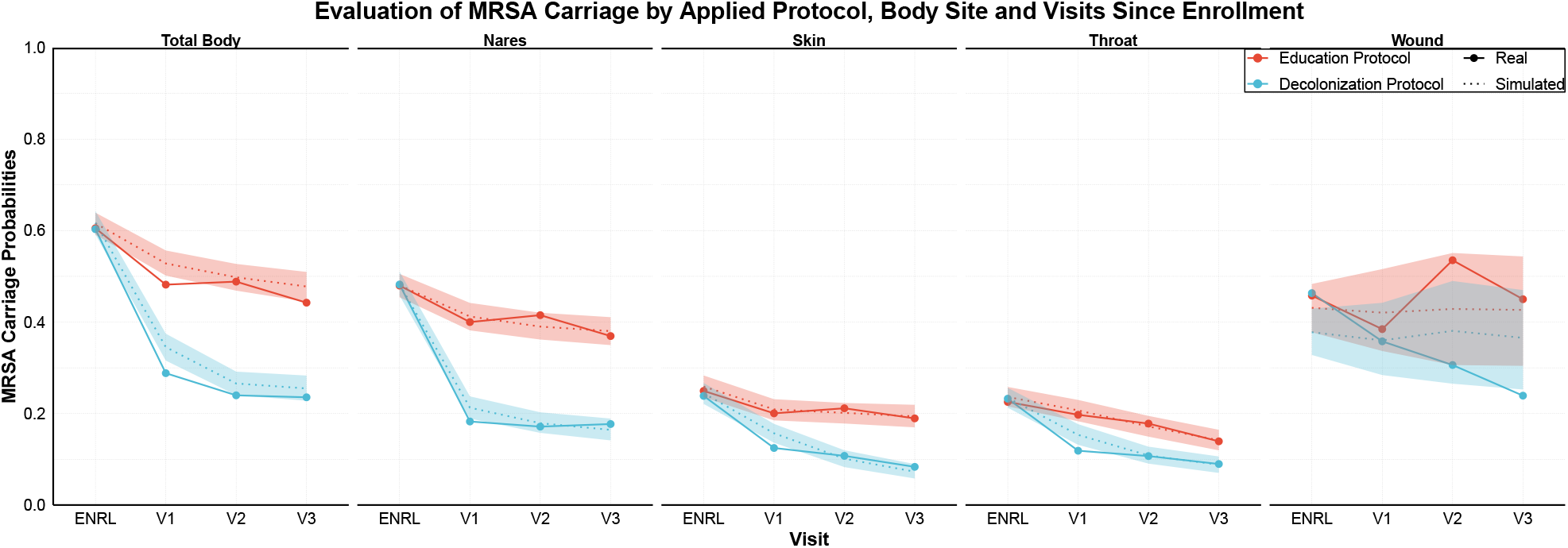
The observed decrease in MRSA carriage over time by body site and study arm compared to the decrease predicted by the model. In the trial, study subjects were in the education group or the decolonization group, which consisted of applying mupirocin to the nares, chlorhexidine mouthwash (CHG Oral) to the throat, and chlorhexidine body washes (CHG Skin) to the skin and, if present, wound. The figure shows site-specific and overall clearance rates in the two groups, along with model predictions. Visits approximately took place in 1 (V1), 3 (V2), and 6 (V3) months after enrollment (ENRL) in the trial. Dotted lines and shaded regions represent the mean and 90% credible intervals (CI) of the simulations, and the solid lines represent values observed in the data.

### The decolonization protocol decreased the persistence of MRSA colonization in the nares and throat independently of other sites

First, we wanted to study the impact of the full decolonization protocol on each particular site while not accounting for any dependencies of carriage between sites (i.e., sites were evaluated in isolation). To do this, we estimated the persistence of colonization for each body site by calculating the probability that the site would be colonized in the next time step, given it was colonized in the previous time step and other sites were not colonized. We found that decolonization protocol significantly reduced the persistence at the nares from 35% (26%-45%) to 10% (7%-14%), and throat from 75% (64%-84%) to 47% (35%-58%), while the persistence of colonization at skin and wound were not affected when these sites were evaluated in isolation (see Figure 3a) [21]. We also estimated the relapse probability, i.e., the probability of a site getting colonized given it was not colonized in the previous time step and other sites also were not colonized. These probabilities were small and did not seem affected by the decolonization protocol (see Figure 3b). We note that the number of samples from wounds was relatively small, which yielded larger uncertainty in the wound-associated parameters. Repeating the analysis only on patients who reported adherence to the protocol led to almost identical results since most (70%) of the data were collected from adherent patients (see Supplementary Figure 5). The model can also estimate other valuable parameters, for example, the sensitivity and specificity of the detection of MRSA in a swab, as those are immediately available as the emission parameters of the CHMM (see Supplementary Figure 3).

**Figure 3.**
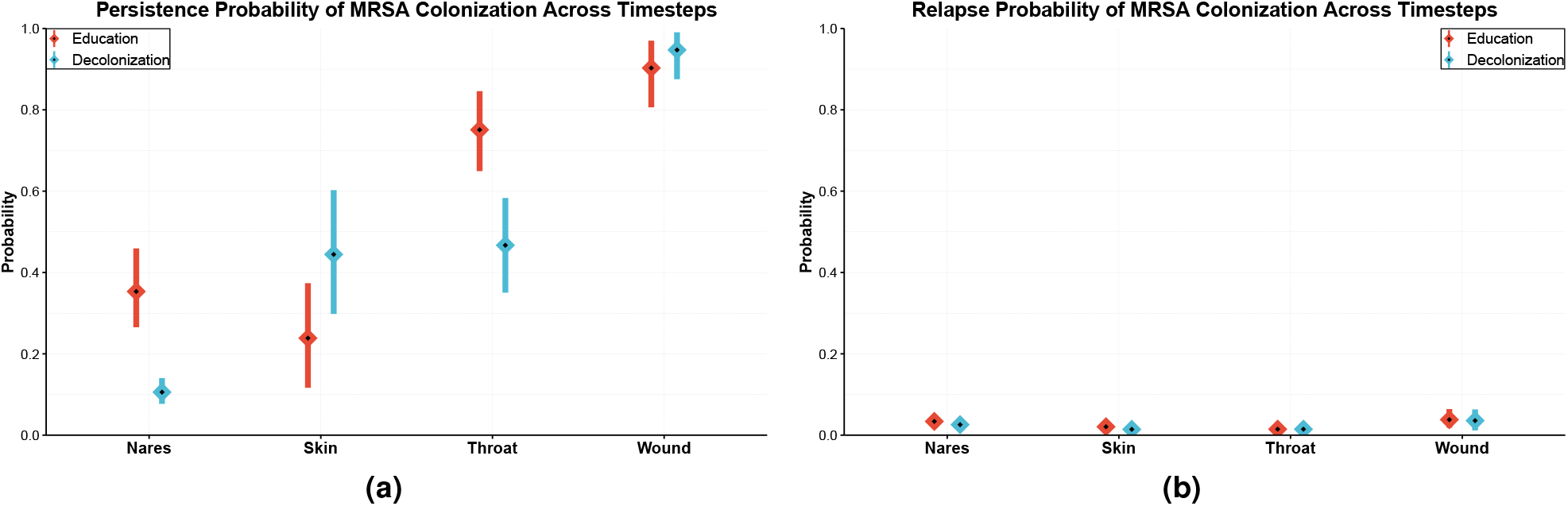
Estimated (a) persistence and (b) relapse probabilities of MRSA colonization by body site. **(a)** *Persistence probability* is defined as the probability of a site will be colonized in the next time step, given it was colonized in the previous time step while other sites were not colonized. **(b)** *Relapse probability* is defined as the probability of a site will be colonized in the next time step, given it was not colonized in the previous time step while other sites also were not colonized. The calculation of the posterior distributions is explained in the Methods. Means and 90% credible intervals (CI) are represented in the figure by diamonds and lines, respectively.

### The decolonization protocol reduced the transmission of MRSA between body sites

We quantified the interactions between body sites in two ways: First, we estimated the probability of MRSA transmission within a time step (corresponding to one month) from the source site to the target (see Supplementary Figure 4) conditional on only the source being colonized, by simulating 1000 datasets from the estimated model. Second, we estimated the proportion of patients with a given MRSA transmission between body sites (see Figure 4), which scales the former transmission probabilities with the observed proportion of patients colonized in the source site of the transmission. First, we found that the most critical determinant of colonization at any site was whether the site itself was colonized in the previous time step. Second, the nares acted both as a source and a sink for transmission, highlighting its role as a hub for MRSA colonization. Third, the strength of most dependencies decreased significantly by the decolonization protocol. These findings, therefore, indicate two mechanisms responsible for the efficiency of the decolonization protocol: 1) the protocol decreased persistence in the nares, which is a central accumulation hub for colonization (see Figure 3), and 2) the protocol weakened links between body sites, reducing self-inoculation (see Figure 4).

**Figure 4.**
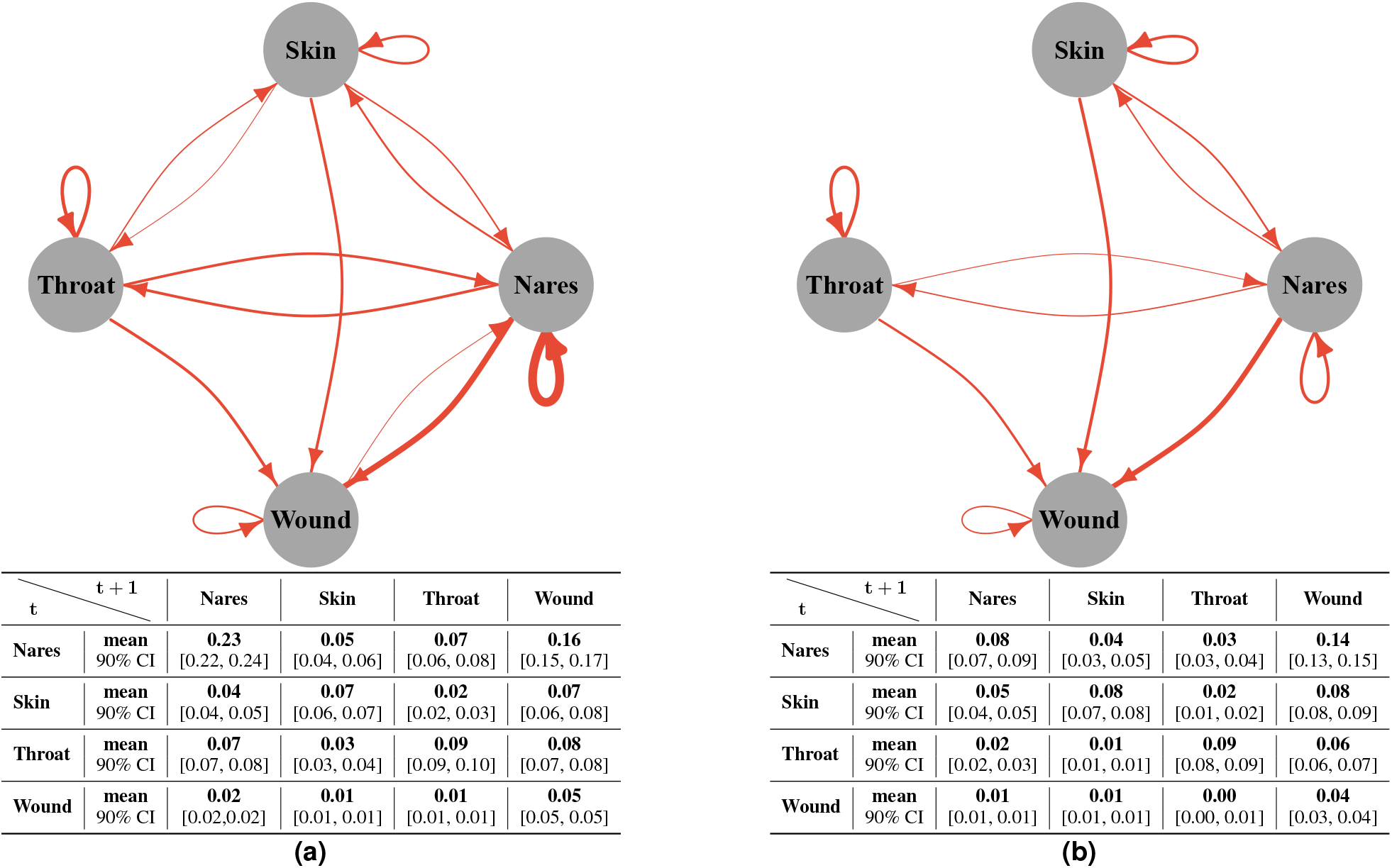
The amount of MRSA transmission among body sites in (a) education and (b) decolonization groups. The edges in the graphs show the estimated proportion of patients with the given transmission between body sites in a time step (corresponding to one month). They are estimated by scaling the transmission probabilities (see Supplementary Figure 4) with the observed proportion of patients colonized in the source site of the transmission. Edges were excluded from the graph if the expected proportion was lower than 0.02. The edge thickness represents the expected value, and the tables show the means and the respective 90% CIs for all relations.

### Nasal mupirocin on the nares was the single most efficient therapy, but all therapies contributed to the efficiency of the decolonization protocol

The importance of the nares has been noted in a study in an intensive care unit [22]. However, the added value of decolonization efforts that focus on body sites other than the nares is not well understood. To quantify the impact of decolonization efforts at each individual site while accounting for interactions between body sites, we predicted the efficiency of hypothetical simplified protocols, consisting of only a subset of the therapies in the full decolonization protocol. Application of nasal mupirocin on the nares alone decreased the estimated total body carriage of MRSA from 61% to 38% (35%-41%), compared to 25% (22%-28%) of the full decolonization protocol and 47% (44%-50%) of education protocol (see Figure 5). Other single therapies were inferred to be less effective than mupirocin, such that only using chlorhexidine mouthwash on the throat (CHG Oral) reduced total body carriage to 42% (39%-44%) and only using chlorhexidine bodywash on the skin and wounds (CHG Skin) reduced total body carriage to 45% (42%-48%), a minor improvement compared to the 47% (44%-50%) of education protocol alone. When we inferred the anticipated success of combinations of two therapies, the best combinations always included mupirocin: CHG Skin + mupirocin decreased carriage to 34% (31%-37%) and CHG Oral + mupirocin to 33% (30%-36%). In contrast, the combination CHG Skin + CHG Oral, i.e., leaving out mupirocin, decreased carriage only to 39%(36%-42%). These results show that all combinations of two therapies had lower efficiency than the full protocol indicating each therapy contributed to the full effect, even if no effect was seen on targeted sites when sites were evaluated in isolation (see Figure 3a). So, even though CHG skin does not appear to affect the target site directly, it is still beneficial, and we hypothesize that the impact comes instead through reduced transmission between body sites (see Figure 4).Figure 6 shows 1) the incremental gain of adding therapies to the protocol and 2) marginal effects of each therapy on the efficiency of the decolonization protocol. Results also indicate that the full effect is greater than the sum of marginal effects, demonstrating the synergy of applying therapies in combination.

**Figure 5.**
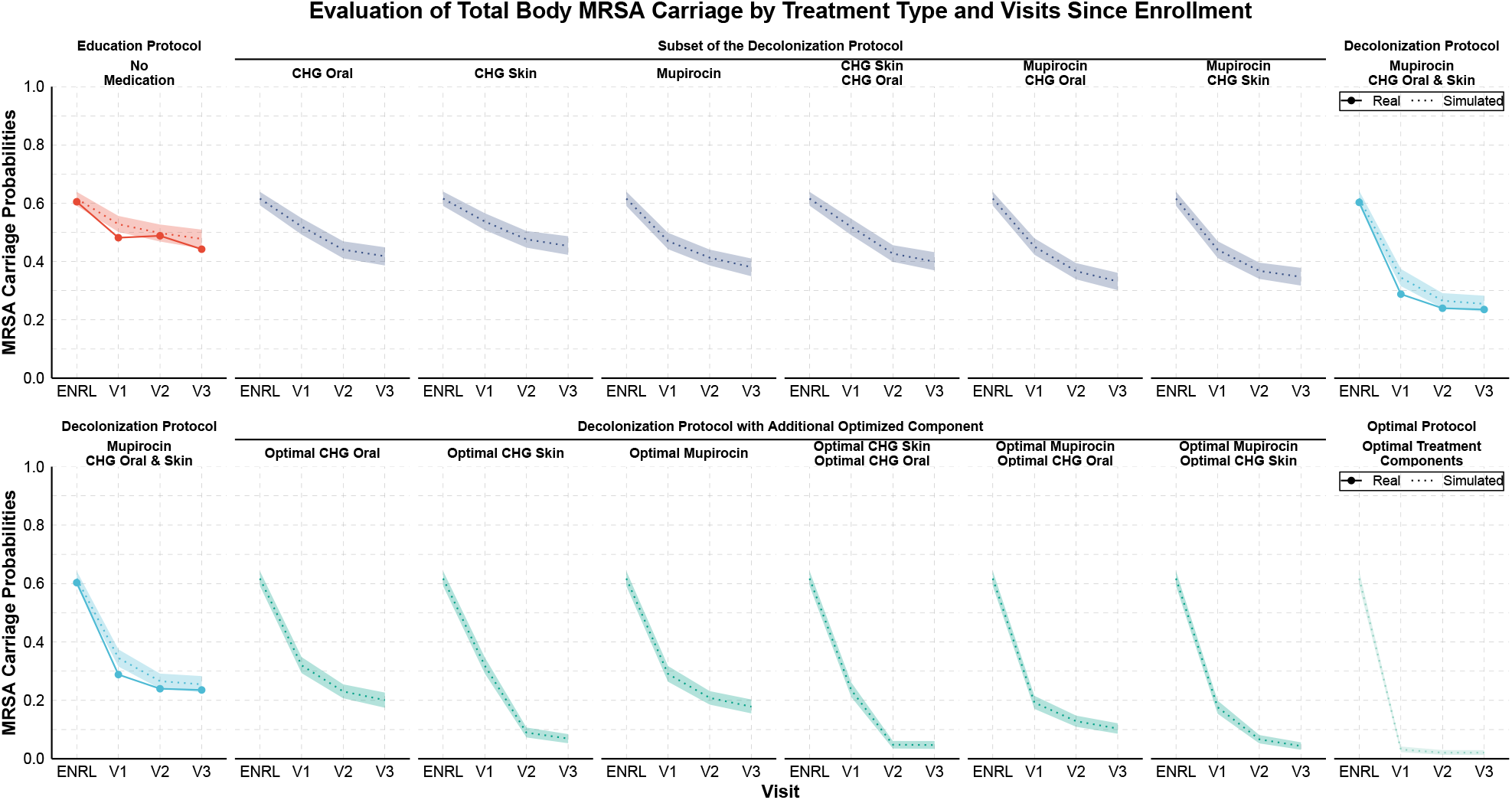
The observed decrease in MRSA carriage over time compared to the decrease predicted by the model for actual and hypothetical therapies. In the trial, study subjects were in the education group or the decolonization group, which consisted of applying mupirocin to the nares, chlorhexidine mouthwash (CHG Oral) to the throat, and chlorhexidine bodywash (CHG Skin) to the skin and, if present, wounds. The predictions for hypothetical therapies assumed either that only a subset of medications was applied or that the decolonization protocol was used with an additional optimized intervention that resulted in immediate clearance of the corresponding target site. Visits approximately took place in 1 (V1), 3 (V2), and 6 (V3) months after enrollment (ENRL) in the trial. Dotted lines and shaded regions represent the mean and 90% credible intervals (CI) of simulations, and the solid lines represent values observed in the data.

**Figure 6.**
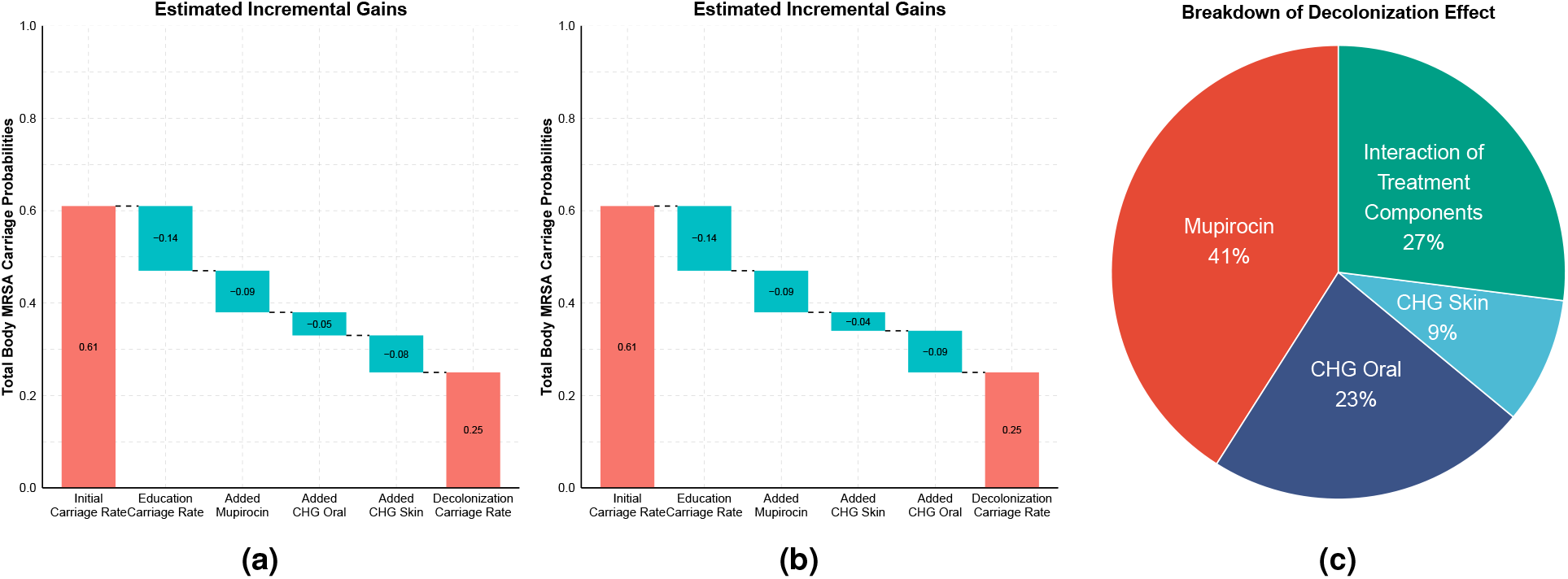
The estimated (a, b) incremental and (c) marginal contribution of each therapy on the efficiency of a decolonization protocol. In the trial, study subjects were in the education group or the decolonization group, which consisted of applying mupirocin to the nares, chlorhexidine mouthwash (CHG Oral) to the throat, and chlorhexidine bodywash (CHG Skin) to the skin and, if present, wounds. **(a, b)** The plots show how the total body carriage at the end of the study decreases when different components are added to the protocol. The cumulative effect is shown for two different orders of adding components. **(a)** In the optimal order (where always the component that most increases the efficiency is added) has CHG Oral added before CHG Skin. **(b)** The order of adding these two components is reversed to show that the most recently added component seems better than it is because it creates an interaction effect between the added therapy and the therapies already included in the protocol. **(c)** The marginal effect of each therapy is calculated using single site therapies on top of education protocol. Interaction of protocol components reflects additional gains achieved in the full decolonization protocol which is not visible in the sum of marginal effects. The confidence intervals are already shown in Figure 5 and written in the text.

### Enhancing clearance at the skin was predicted to achieve the most significant gain in overall decolonization success

The key metric of success for decolonization is the extent to which it reduces MRSA carriage. We used our model to predict the impact of enhanced effects on the components of the decolonization protocol. We assumed optimal effects that resulted in immediate clearance of a specific site. This could be achieved either through a more effective replacement product, an additional product applied to the site on top of the existing protocol, or training to achieve the appropriate application of the protocol with high adherence leading to immediate clearance. The result reflected the relative inefficiency of the original CHG Skin therapy (see Figure 3a and Figure 5), such that the most significant improvements were seen by improving this therapy. In particular, the estimated final carriage with the full protocol but optimized nasal mupirocin was 17% (15%-20%), while optimizing the throat (CHG Oral) or skin/wound (CHG Skin) therapies decreased the carriage to 20% (17%-22%) or 6% (5%-8%), respectively. Optimizing therapies both on nares + the throat has the potential to reduce carriage further down to 10% (8%-12%) compared to 4% (3%-5%) of the nares + skin/wound, and 4% (3%-6%) of the throat + skin/wound.

## 3 Discussion

We presented a model for the dynamic process of MRSA carriage that quantified interactions among colonization at different body sites, quantified the impact of each therapy, and predicted the decrease in MRSA carriage over the study period. We found that not all therapies contributed equally to successful decolonization. Among the CLEAR Trial components, mupirocin was estimated as the most effective therapy, but our results also indicated that all therapies were essential to the full effect of the complete protocol. Furthermore, our results indicated that enhancing the chlorhexidine bodywash therapy has the greatest potential for improving the effectiveness of the decolonization regimen.

As with all modeling, our analysis made several simplifying assumptions. First, the analysis focused on MRSA clearance on individuals, but the decolonization protocol may have other benefits, e.g., reduction in transmission between patients [23, 24]. Second, we assumed that the missingness of observations (including trial exits, skipped visit or non-present wounds) did not depend on the colonization status of the body site, which is likely not always correct; for example, if a previously colonized wound is healed and not colonized anymore, no additional samples would be taken from the wound. Finally, in our optimal therapy analysis, we assumed the sensitivity and specificity of the swabs were not affected by the new optimal therapy. However, the importance of the therapies on these parameters has been noted in a previous study [25]. The model could be improved further by 1) incorporating the type and the burden of MRSA because heterogeneity in treatment responses among patients and strain types of MRSA might affect the decolonization dynamics [26], and 2) using semi-Markov approaches in the CHMM to explicitly model the variations in time intervals between the visits (currently the longer intervals were assumed to include missing in-between observations). Our analysis was based on a Bayesian approach, requiring a specification of priors on model parameters to express what is believed about their values before seeing the data. We used weakly informative priors in accordance with the literature [25], and comparison included in the Supplementary Table 1 shows that the model fit was not sensitive to the exact values of the priors. Despite the simplifications, the method successfully passed a comprehensive set of diagnostic tests for model fit and convergence of the algorithm.

This study indicated that the impact of individual interacting components of complex clinical protocols and RCT data can be rigorously deconvoluted and assessed *in silico* by machine learning tools like those used here, thereby enabling the design of interventions that are more efficient, easier to adhere to, and more likely to be successful. This analytical approach thus demonstrates how data from randomized controlled trials can inform about biological processes as well as guide improvements in clinical protocols and decision-making.

## 4 Methods

### Isolate Collection

MRSA isolates were collected as part of the CLEAR (Changing Lives by Eradicating Antibiotic Resistance) Trial. The trial was designed to compare the impact of a repeated decolonization protocol plus education on general hygiene and environmental cleaning to education alone on MRSA infection and hospitalization [3]. Study subjects in the trial were recruited from hospitalized patients based on an MRSA positive culture or surveillance swabs. After recruitment, swabs were obtained from different body parts of subjects (nares, skin, throat, and wound) around the time of hospital discharge (ENRL) and at 1,3,6 and 9 months (V1-V4, respectively) following the initial visit. The application of decolonization protocol lasted only for 6 months, and consequently, we only modeled visits until V3 in this study. We note that some enrolled study subjects, despite a positive culture (clinical or surveillance) during the hospital stay, did not have discharge swabs positive for MRSA at the first time point (ENRL). More details about sample collection and data preprocessing can be found in the reference [3].

### Model

In this section, to describe the properties of CHMM, we initially described the properties of HMMs. Then, we presented the details of CHMMs, which were built on the HMM.

### Hidden Markov Model

Markov models are a well-known tool for time series analysis. Hidden Markov models (HMM) are a special case where Markov process is observed indirectly through noisy observations. Discrete-time discrete-state hidden Markov model with a latent sequence ***π***, *π*_*t*_ ∈ {1, …, K} and observations **x**, *x*_*t*_ ∈ {1, …, L} for *t* ∈ {1, …, *τ*}, is defined as

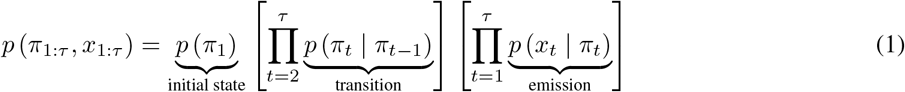

where K is number of latent states, *τ* is the number of time steps, and *L* the number of possible observed states. Parameters of the model are the initial state probability *π*_0_, transition probability *T*, and emission probability *E*, defined as

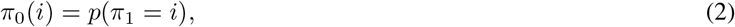

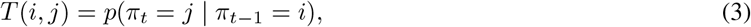

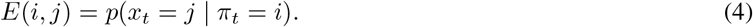

Note that the rows of *T* and *E* are probability distributions by definition. We will jointly denote the model parameters as *θ* ={*π*_0_, *T, E*}. If *θ* is known, then we can estimate the latent states *π* conditional on parameters *θ* and observations **x** using the forward-filtering backward-sampling algorithm.

### Forward-Filtering Backward-Sampling

Filtering is the estimation of the current hidden state by using all observations so far, *p* (*π*_*t*_ | *x*_1:*t*_) [19]. This is known as *forward-filtering* and it can be represented as follows:

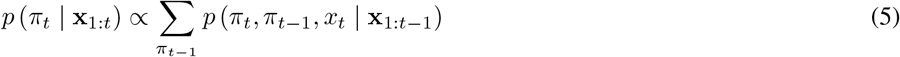

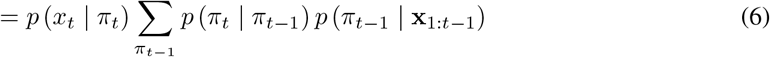

We define *α* (*π*_*t*_) = *p* (*π*_*t*_ |**x**_1:*t*_) and substitute that into Equation 6, which yields the following recursive equation known as *α-recursion* or *forward recursion*;

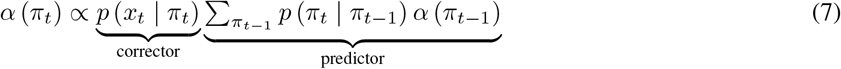

The recursion starts with *α* (*π*_1_) = *p* (*π*_1_ |*x*_1_) = *p* (*x*_1_ |*π*_1_) *p* (*π*_1_) */p* (*x*_1_). This shows that the filtered distribution *α*(.) is propagated through to the next time-step, where it acts like a **‘prior’**. In other words, at each time step the calculated posterior becomes the new prior for the next time-step [27].

To estimate the posterior distribution of latent states, we sample from the joint distribution of the latent sequence, *p*(***π***_1:*τ*_ | **x**_1:*τ*_):

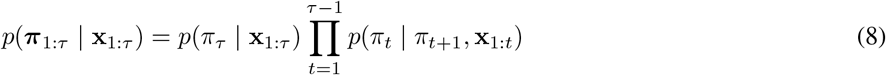

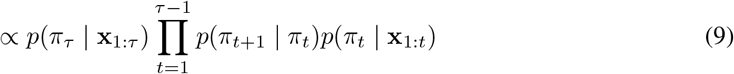

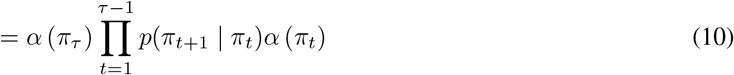

According to Equation 8, to be able to compute the posterior distribution of latent states, we need **‘time-reversed’** transitions *p*(*π*_*t*_ |*π*_*t*+1_, *x*_1:*t*_), which are obtained using *α-recursion*s from the forward filtering. The simulation starts from the end of the sequence and proceeds backwards recursively. Starting point is

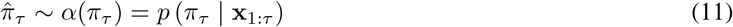

For each time step *t*, the unnormalized sampling probability can be calculated using the previously sampled latent state 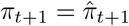, and the 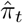 can be sampled using following:

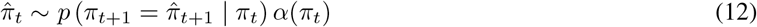

This procedure is known as forward-filtering backward-sampling [27].

### Learning Model Parameters

Conditionally on the latent states and observations the parameters *θ* = {*π*_0_, *T, E*} of the model can be estimated. The posterior distribution 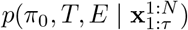 is given by

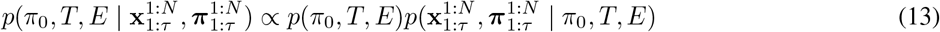

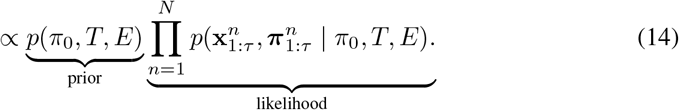

Factorizing the likelihood term in Equation 14 as in Equation 1, we can get the following result:

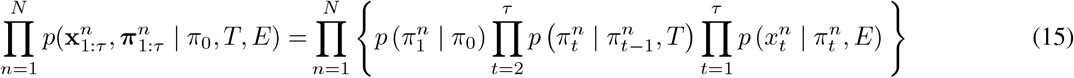

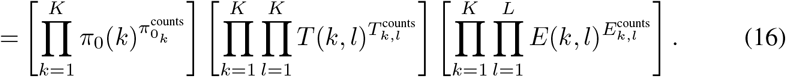

Since, *π*_0_, rows of the *T*, and rows of the *E* are probability distribution, [·]’ s inside the Equation 16 corresponds to Multinomial distribution with parameters 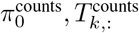 and 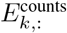, where 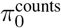 corresponds to the initial state counts, *T* ^counts^ corresponds to the state to state transition counts, and *E*^counts^ corresponds to the state to observation emission counts. Since the Dirichlet is conjugate prior for the Multinomial distribution, we set a Dirichlet prior for *θ* = {*π*_0_, *T, E*} with hyperparameters 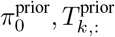, and 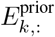, respectively. Because of the conjugacy, the resulting posterior in Equation 14 is also a Dirichlet distribution:

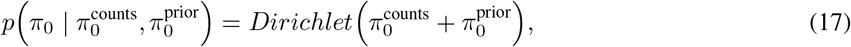

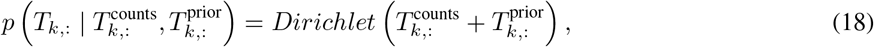

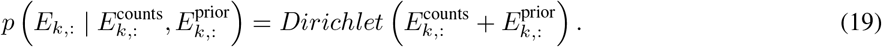

### Inference of HMM with MCMC Sampling

There are several ways to estimate the parameters of HMM when both hidden states and model parameters *θ* are unknown, for example the expectation-maximization algorithm, variational Bayes, or MCMC sampling. In this paper, we will use MCMC. The goal of MCMC is to produce draws from the posterior *p*(***π***, *θ* | **x**). The Gibbs sampler is an MCMC algorithm suitable for high dimensional problems, such as the HMM, and it samples parameters one-by-one using their distributions conditional on the other parameters. In the HMM, it will alternate between sampling the model parameters *θ* conditional on the latent sequence ***π*** from *p*(*θ* | ***π*, x**), and the latent sequence ***π*** conditional on the model parameters *θ* from *p*(***π*** | *θ*, **x**). The algorithm iterates the following steps *N* times, which gives *N* draws from the posterior:

1. Sample the model parameters 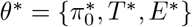 from Equation 17, Equation 18, and Equation 19, respectively. (Note that they are independent of each other given latent sequence.)
2. Sample the latent sequence of HMM using the updated model parameters *θ* using the Forward-Filtering Backward-Sampling algorithm.

Before calculating the posteriors, the first *N*_0_ samples are discarded as a convergence (warm-up) period of the Markov chain. The plate diagram of the HMM is given in the Figure 7a, and the algorithm is described in Algorithm 1.

#### Algorithm 1 Hidden Markov Model

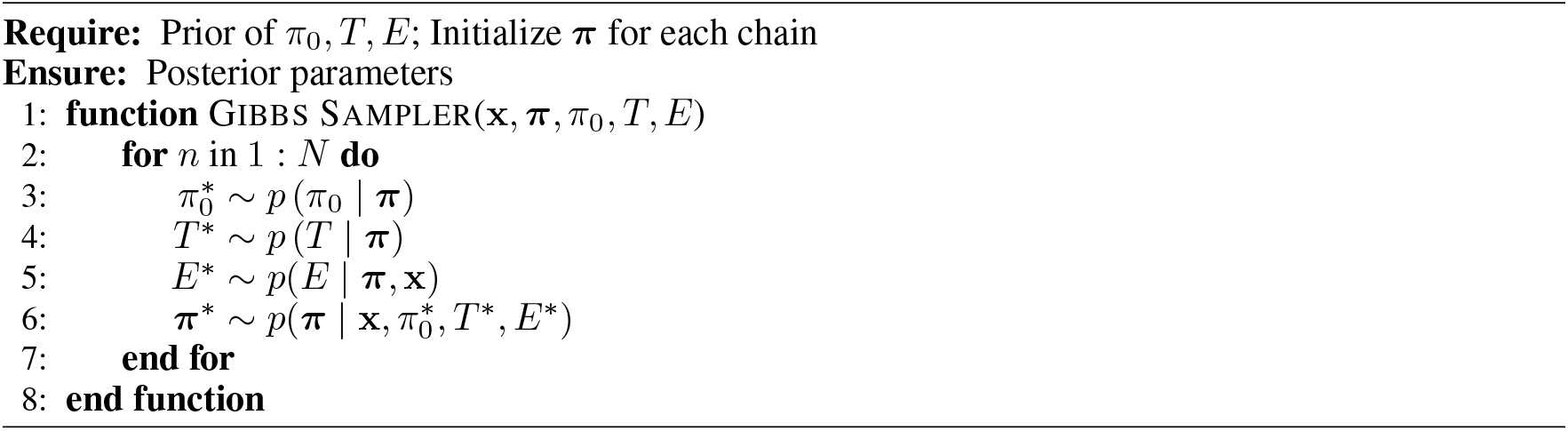

**Figure 7.**
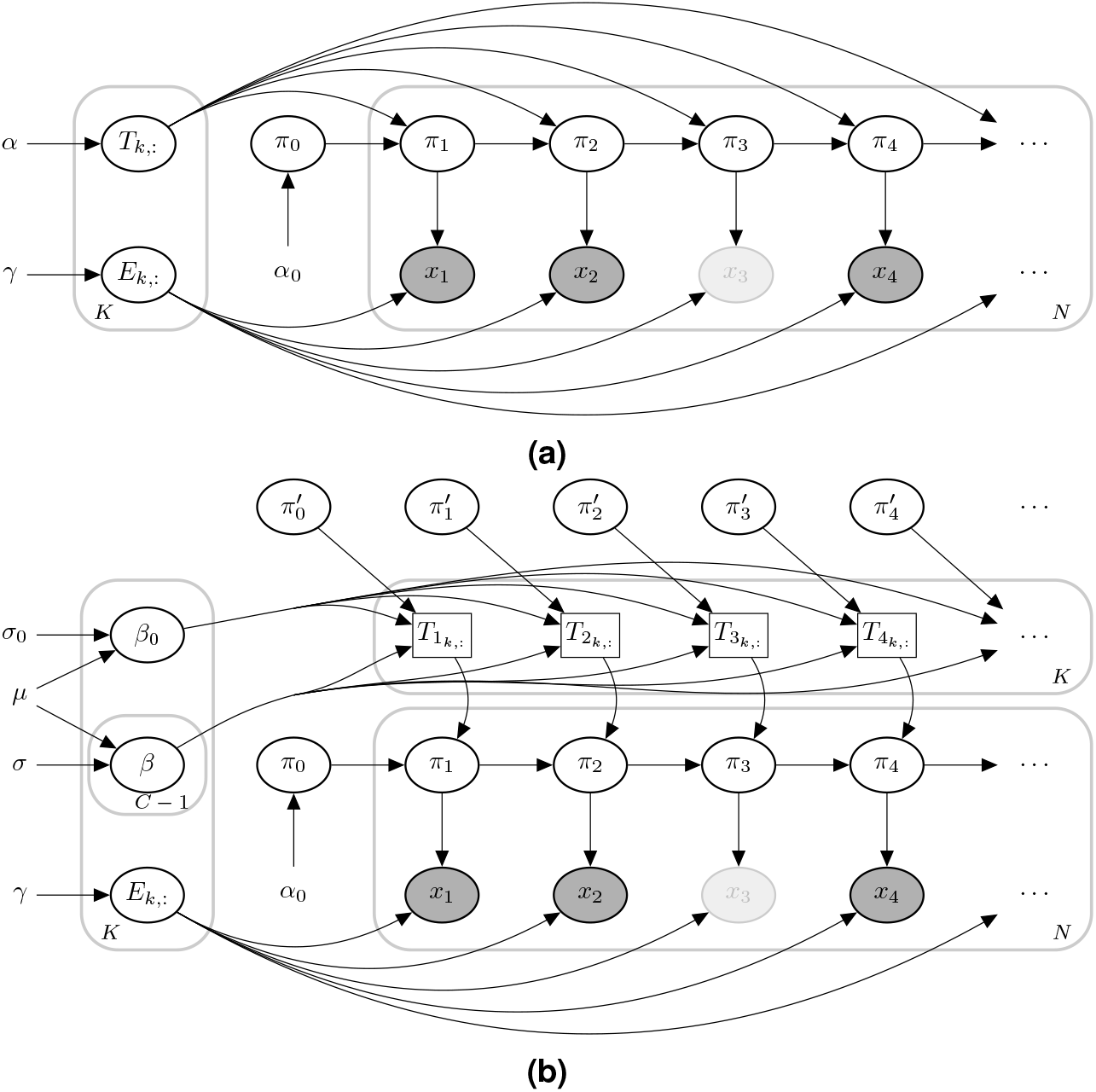
Plate diagram of (a) hidden Markov model (HMM) and (b) coupled hidden Markov model (CHMM). Here, for clarity, we illustrate only one chain. *π*_*t*_ represents the unobserved true states, and *x*_*t*_ represents the noisy observations at time *t*. The faded observation nodes correspond to missing values. Plain parameters are hyperparameters of the model. Edges from *π*_*t*_ to *x*_*t*_ represent *emission probabilities*. In the CHMM, the next state in one site depends on the previous states of the other sites. It is illustrated by *π*^′^. Therefore, *transition probabilities* change at each time step.

### Coupled Hidden Markov Model

In the set of ordinary HMMs, transition parameters do not change within the chain. However, in the CHMM, transitions in one chain are affected by other chains. In principle, it would be possible to define a single joint HMM where the latent state represents the latent states of all individual chains jointly and adapt the solution for the HMM described above. However, this is inefficient when there are many chains because the number of states would grow as *O*(*K*^*C*^), where *K* is the number of hidden states in a chain and *C* is the number of chains. On the high level, our key idea is that the transition matrix of each chain is modeled conditionally on the states of the other chains (and not as a single large joint transition matrix). A similar but simpler formulation was considered by reference [12]. Note that a transition matrix *T* for a specific chain will not be time-independent anymore, instead, changes at each time step depending on the states of the other chains. The graphical representation of the model (showing just two chains) is given in Figure 1, and the plate diagram of the CHMM is given in Figure 7b. There are *O*(*KC*) parameters in this formulation, which is much more efficient for the increased number of chains. In theory, CHMM is a low-rank estimation of one joint HMM.

Defining the transition matrix is a critical part of the algorithm, and it is essential to carry as much information about the other chains as possible. We model the dependencies between the chains with parameters ***β***, where each *β*. is a matrix of the same size with the transition matrix. Assume there are *C* chains and let ℂ denote the set of chains. Further, assume that each chain can be in one of *K* possible states, and let 𝕂 denote the set of states. Finally, assume that there is a baseline state 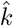such that if a chain is in that state, it does not affect other chains (in our application, this state corresponds to the absence of MRSA colonization in the respective body site). The transition matrix for the chain ĉ, at time *t*, denoted as 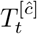, is defined by;

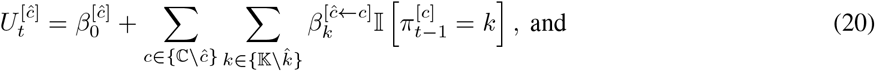

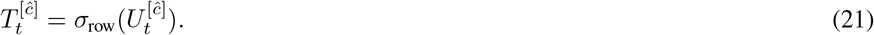

The transition matrix 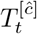 is obtained in Equation 21 by applying a row-wise softmax operator *σ*_row_ to the unnormalized transition matrix 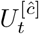. Parameter 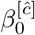 corresponds to an intercept matrix and it specifies the transition matrix of the target chain ĉ when all other chains are in the baseline state 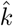. Parameter 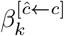 represents the impact of chain *c* on the target chain ĉ and it is added to 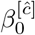 whenever chain *c* was in state 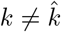 in the previous time step. We will denote all the parameters for target chain ĉ as ***β***^[ĉ]^.

In Equation 20, the unnormalized transition probability *U* is an additive function of the latent states of the other chains; hence we call it the *Additive-CHMM*. We also implemented a simple formulation than the Additive-CHMM, which we call *Or-CHMM*. In the Or-CHMM, the target chain is not affected by the other chains individually; instead, there the target chain is affected whenever *any* of the other chains is in some state different from the baseline state, and the unnormalized transition matrix is given by:

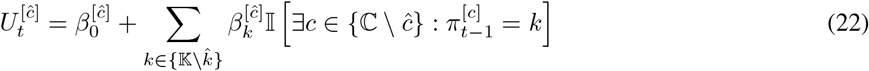

In our application we have *K* = 2, corresponding to the presence and absence of colonization in a given body site. In this case, Equation 22 has a simple form: If all other chains are in state 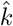, the output is only 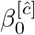, and if any of the other chains is in a state other 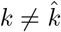, then the output is the sum of 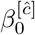 and 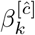.

### Design and Interpretation of the *β* Parameters

In the CHMM, the transition matrix *T* for each chain is modeled as a function of *β* parameters, where each *β*_*k*_ is a matrix of the same size as the transition matrix. Therefore, the *β* parameters are a list of matrices. Since the rows of a transition matrix *T* are assumed independent, we also model *β* parameters such that the rows of those matrices are independent. However, to avoid redundant parameters, the rows on each row of *β* are assumed to sum to zero. In practice, we sample the first *K* − 1 parameter on a given row and set the *K* th element to equal the negative of the sum of all the other parameters. This constraint ensures that the mapping from *β* to *T* is one-to-one.

To give an interpretation to the parameters *β* in the Additive-CHMM, we start with Equation 21 and write the softmax for a single element of a transition matrix *T* (*i, j*):

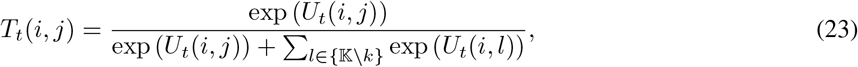

from which it follows after straightforward algebra:

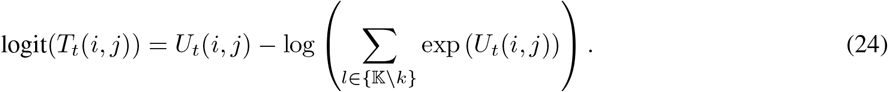

In our application *K* = 2 and sums of the rows of *β*_*k*_ are assumed equal to 0, so consequently also the rows in the *U*_*t*_ sum to zero, which gives us:

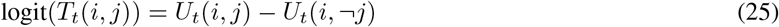

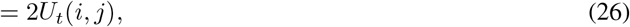

Where ¬ *j* refers to the other element on the row that is not *j*. Therefore, in this 2-dimensional case, all *β*_0_ values correspond to half of the log-odds of the respective transition probability, and similarly, parameters *β*_*k*_ representing the interactions between the chains correspond to half of the change in the log-odds because of the presence of colonization in the other chain.

### Adaptive Metropolis-Hastings Within Gibbs Sampling

So far, we have described the design specifications of the CHMM. Since transitions are changing at each time step, it is not feasible to calculate their sufficient statistics as in Equation 16. To resolve this, we sample the *β*^[ĉ]*^ using a Metropolis-Hastings(MH) step within the Gibbs sampler because conditional on the latent states of all the chains, the transition parameters are fully determined by the *β*^[ĉ]^ parameters. To get a draw from 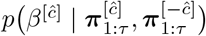, where 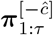 states of the other chains, we use the following steps;

1. Make a proposal *β*^[ĉ]*^ ∼ *q*(*β*^[ĉ]^)
2. Transform *β*^[ĉ]*^ samples to transition probabilities 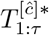 according to latent states of the other chains, *π*^*′*^
3. Accept the proposal *β*^[ĉ]*^ with probability

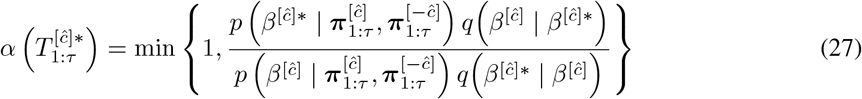

where

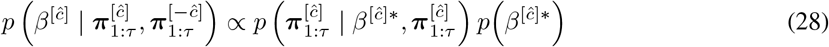

The first quantity on the right-hand side of the Equation 28 can be directly calculated with the transition parameters 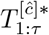 for corresponding chain. We used the Gaussian proposal in which the previously sampled *β*^[ĉ]^ is the mean such that *q* (*β*^[ĉ]*^ | *β*^[ĉ]^) = *N* (*β*^[ĉ]*^ | *β*^[ĉ]^, *c*^*2*^ Σ). As a variance parameter, we used a fixed diagonal covariance matrix during the warm-up period and then used the covariance matrix estimated from the previous samples. We initially set 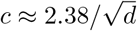 as Metropolis jumping scaling factor since it is theoretically the most efficient scaling factor [28], where *d* is the dimension of the sampling. Then we adaptively scaled the scaling factor *c* as described in the reference [29]. Since this proposal distribution is symmetric (i.e., Normal), *q β*^[ĉ]^ *β*^[ĉ]*^ and *q β*^[ĉ]*^ *β*^[ĉ]^ in Equation 27, cancel out each other.

To sample the latent sequences we need to modify the Forward-Filtering Backward-Sampling algorithm [15], since we need to draw samples from 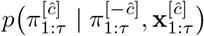 instead of 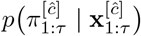 The modified *forward-filtering* is defined as follows:

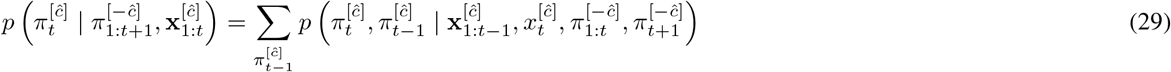

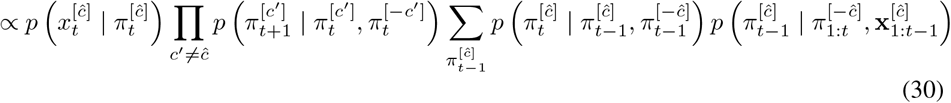

If we define 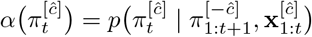 and substitute that into Equation 30, we can acquire *modified α-recursion*;

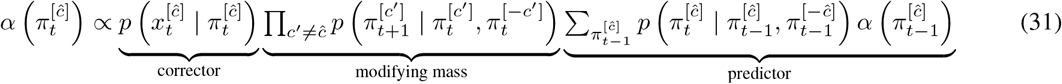

Respectively, the posterior distribution of the latent sequences for each chain can be acquired as follows:

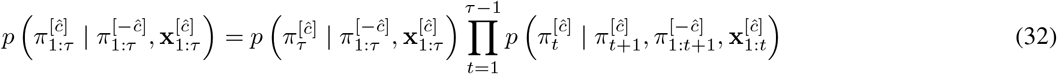

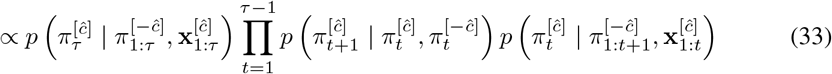

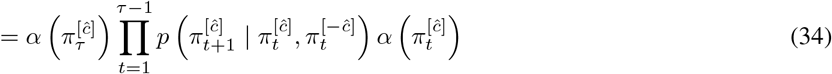

Therefore we can use the sampling procedure described in Equation 11 and Equation 12. Algorithm 2 summarizes the whole algorithm. To validate the implementation, we used the algorithm to estimate parameters in simulated data. The results in Supplementary Figure 2 show that the algorithm estimated the parameters correctly and yielded well-calibrated posterior distributions.

#### Algorithm 2 Coupled Hidden Markov Model

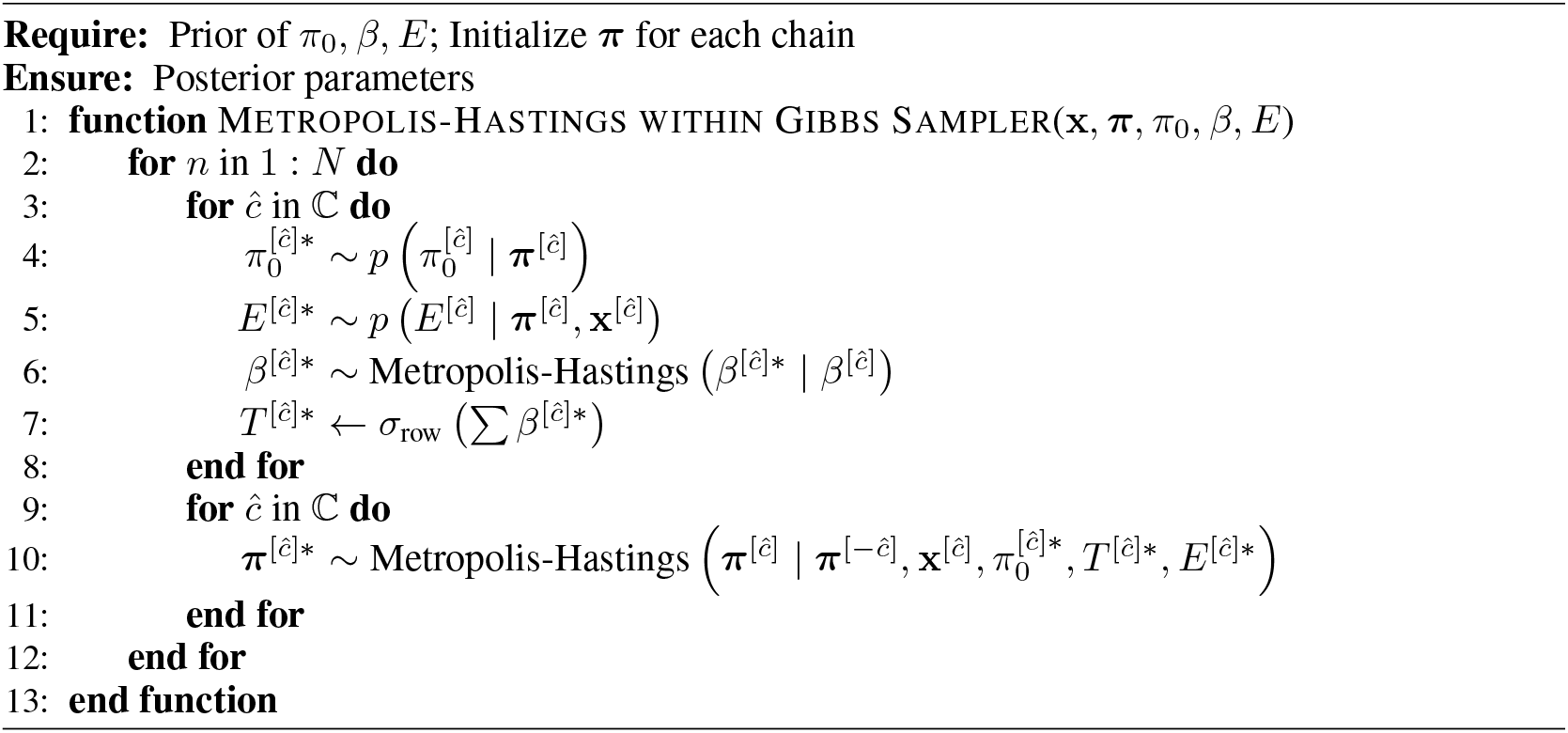

### Implementation details

We set the initial covariance matrix for the Metropolis proposal as 0.01 × *I*, where *I* is the identity matrix, which corresponds to a step size giving the optimal acceptance rate of ≈ 23% [30]. We set the prior of *β*_0_ as *N* (*β*_0_ | 0, 1), which is almost uninformative so that the estimates are not affected strongly by the prior. For the rest of the *β* parameters, denoted by *β*_*k*_, we used sparsity encouraging Horseshoe prior with mean and scale parameters are 0 and 0.25, respectively (see Supplementary Materials). We used a uniform prior on the initial state probabilities *π*_0_, and weak Dirichlet priors for the rows of emission probabilities *E* such that we set the value to 30 for specificity and 15 for sensitivity. We set rest of emission priors to 1, which corresponds to uniform prior. In such a formulation, except for the initialization, the prior has a negligible effect because it is summed with observation counts during inference. We drew 50,000 MCMC samples, and we set the warm-up length as 25,000. Posterior probabilities are calculated using the remaining MCMC samples. HMM and CHMM implicitly assume that time intervals between observations are the same, which is not the case in our data. Therefore, during the model training, we assumed that there are missing observations at 2, 4, and 5 months after enrollment.

## Data Availability

The CLEAR (Changing Lives by Eradicating Antibiotic Resistance) Trial demonstrated that the use of a post-discharge decolonization protocol in MRSA carriers reduces infection and hospitalization rates; ClinicalTrial.gov number NCT01209234, see reference https://doi.org/10.1056/NEJMoa1716771 for informed consent and institutional review board approvals. The data used in the present article is available on request from the corresponding author Susan Huang of the https://doi.org/10.1056/NEJMoa1716771, a co-author of this study.

## Data Availability

The CLEAR (Changing Lives by Eradicating Antibiotic Resistance) Trial demonstrated that the use of a post-discharge decolonization protocol in MRSA carriers reduces infection and hospitalization rates; ClinicalTrials.gov number NCT01209234, see reference [9] for informed consent and institutional review board approvals. The data set used in the present article is available on request from the corresponding author Susan Huang of the reference [9], a co-author of this study.

## Code Availability

The R package is available at https://github.com/onurpoyraz/chmmMCMC.

## Author Contributions

YHG and PM perceived the study. OP, PM, and YHG designed the model. OP implemented the model and run the analyses. OP, SSH, YHG, and PM interpreted the results. MRAS, LGM, JAM, and SSH prepared the data. OP, PM, YHG wrote the manuscript with comments from all authors.

## Supplementary Materials

### The Decision of the Model Used

We developed the CHMM methods to investigate the interactions between the MRSA colonizations at different body sites. Unlike HMM, the probability of MRSA colonization at a given site depends on the MRSA colonization at all body sites in the previous time step. Here, we developed two novel formulations: *Additive-CHMM* and *Or-CHMM*. In the former formulation, the probability of colonization at a given site is an additive function of colonization at the other sites. This idea properly formulates the effect of the other colonized sites on colonization at the target body site. However, it imposes an inductive bias that the target site is affected by those independently when there is more than one colonized body site. In the latter formulation, the probability of colonization in the body site of interest changes if *any* of the other sites is colonized. This formulation uses fewer parameters than *Additive-CHMM*, and semantically it imposes an inductive bias that the location of colonization is not important. Therefore, this formulation does not identify the source of transmission. On the other hand, both CHMM formulations can identify the affected sites. Finally, we also used the site-specific standard HMMs for each site (we call it *Standard-HMMs)* to check whether our CHMM formulations provide a significant improvement over HMM or not. With the standard HMMs, one can not estimate any interaction among colonization at different body sites. We trained all of the models on our dataset, and using the results at Supplementary Table 1, we see that both CHMM methods have a better score than HMM even though they have more parameters. Additionally, the model comparison results at Supplementary Table 1 showed that Additive-CHMM had superior accuracy compared to Or-CHMM or a set of site-specific standard HMMs.

### Model Comparison

We compared three models: the site-specific Standard-HMMs as a baseline (i.e., modeling each site independently with a different HMM), the Or-CHMM, and the Additive-CHMM model using six different initial parameterizations. First, we considered prior *N* (*β*_*k*_| 0, 0.5), as in *β*_0_, and prior *N* (*β*_*k*_ | 0, 0.25), to see how sensitive the performance is to the strength of the prior. In addition to these Gaussian priors, we also considered the Laplace and Horseshoe distributions, which are sparse priors that encourage values close to zero. Note that *β*_0_ always had the same uninformative Gaussian prior because the *β*_0_ values represent the impact of the history of a chain on its future values, which is expected to be non-sparse. On the other hand, *β*_*k*_ values correspond to the effects of the other chains, which may be zero in practice.

We compared the models using the leave-one-out cross-validation (LOO-CV), which measures model performance by estimating pointwise out-of-sample prediction accuracy from a fitted Bayesian model using the log-likelihood evaluated at the posterior simulations of the parameter values [20]. There are two different widely used information criteria (IC): 1) The Akaike Information Criteria (AIC), which is the summation of the negative log-likelihood of the model and the number of parameters, 2) the Watanabe-Akaike Information Criteria (WAIC), also known as the Widely applicable information criteria, which is the generalized version of the AIC onto singular statistical models [31]. LOO-CV is more computationally intensive than AIC and WAIC. Still, it is theoretically preferred over AIC and WAIC for Bayesian models [20]. Therefore, we used LOO-CV as an IC. For this criterion, a lower score is better. Supplementary Table 1 presents all the scores used to evaluate model performance.

**Supplementary Table 1.** Comparison of models using LOO-CV information criterion in education and decolonization groups. All priors are 0 mean, and *σ*’s represent the scale parameters of the distributions. The best formulation is represented by bold text. For LOO-CV information criterion, a lower score is better.

| Group<br>Model<br>Prior | Education Group |  |  | Decolonization Group |  |  |
| --- | --- | --- | --- | --- | --- | --- |
|  | Standard<br>HMMs* | Or<br>CHMM | Additive<br>CHMM | Standard<br>HMMs* | Or<br>CHMM | Additive<br>CHMM |
| Gaussian $\sigma = 0.25^\dagger$ | 10442 | 9265 | **8373** | 8047 | 7064 | **6735** |
| Gaussian $\sigma = 0.5$ | | 9281 | 8538 | | 7070 | 6751 |
| Laplace $\sigma = 0.25$ | | 9274 | 8452 | | 7101 | 6803 |
| Laplace $\sigma = 0.5$ | | 9297 | 8496 | | 7083 | 6806 |
| Horseshoe $\sigma = 0.25$ | | 9279 | 8413 | | 7076 | 6792 |
| Horseshoe $\sigma = 0.5$ | | 9301 | 8441 | | 7081 | 6700 |

**Supplementary Figure 1.**
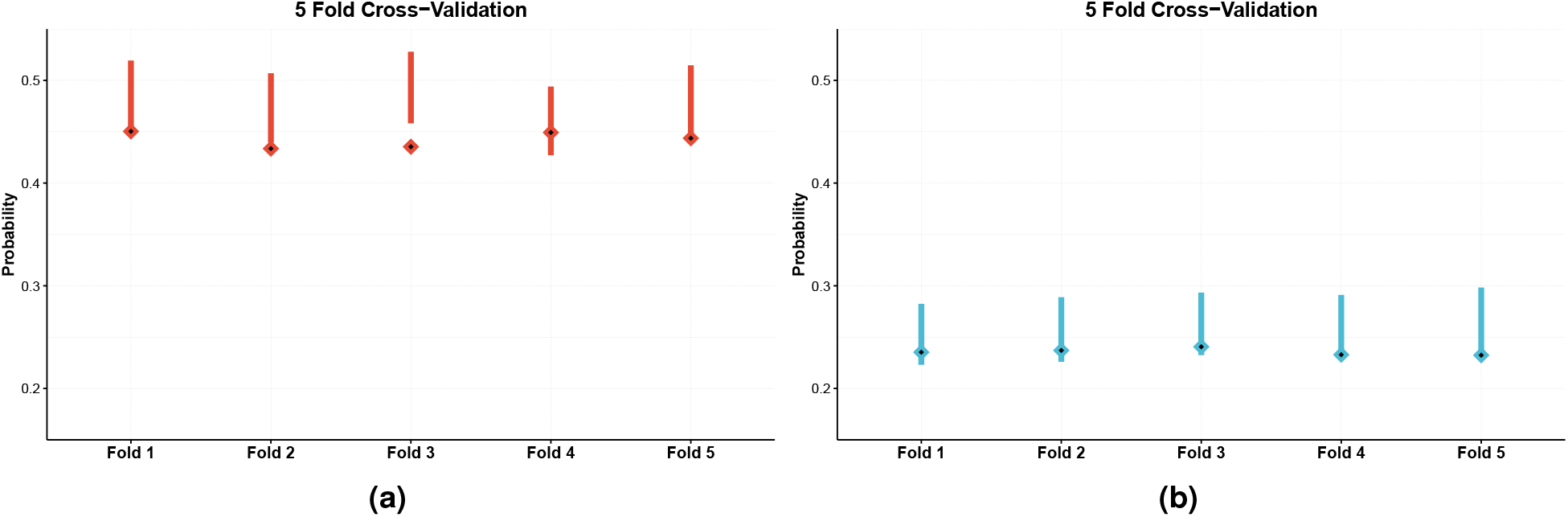
The results of 5-Fold cross-validation on (a) education and (b) decolonization group. **(a)** The model is trained with patients in the train fold (80% of the data, excluding the test fold), and then a prediction is formed about the fraction of patients colonized at the end of the trial (the lines show 90% CIs). The true fraction of the colonized patients in the test fold (20% of the data) is shown with a diamond. We see that in most cases, the 90% CI includes the true value. Also, we see the predicted final fractions seem to be slightly larger than the observed fraction of colonized patients, which means that the predictions are somewhat conservative (rather under-than overestimating the decrease in colonization), which is expected with a finite data set because shrinkage priors are used in the model.

**Supplementary Figure 2.**
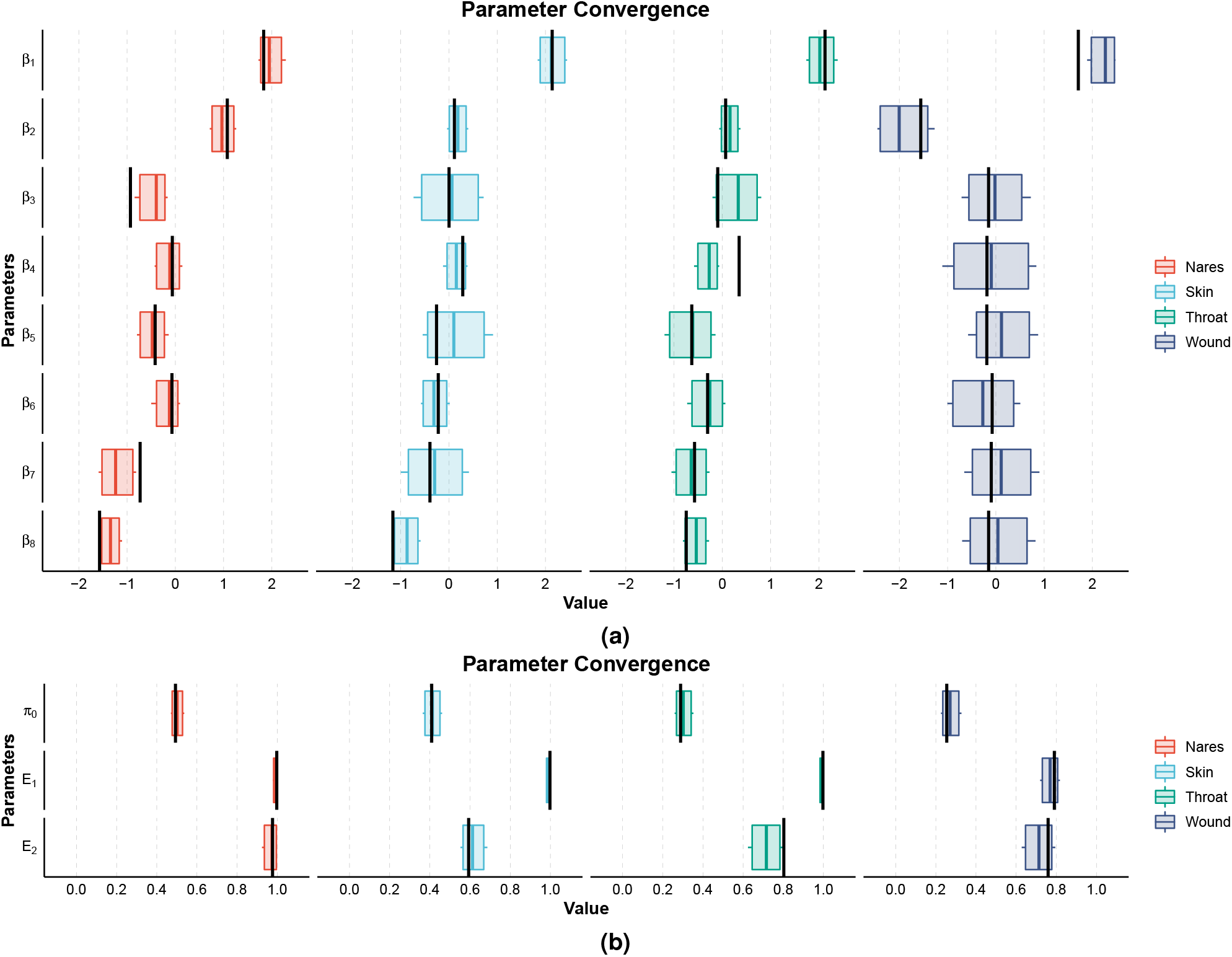
Parameter convergence in the simulated data: **(a)** *β* parameters correspond to change in the log-odds (see Design and Interpretation of the *β* parameters), and they determine the transition probabilities at given site, while **(b)** *π*_0_ and *E* parameters are probability distributions, where *π*_0_ shows the colonization probability for a given chain at hospital discharge and *E*_1_ and *E*_2_ correspond to specificity and sensitivity of the MRSA colonization detection, respectively. To create realistic test data, we used parameters estimated with the real data to simulate the test data. Further, the simulated data mimics the real data in terms of sample size and the pattern of observed samples: There are no samples in the second, fourth, and fifth months (because there were no corresponding visits in the real data), and some samples were not collected because of trial exits, skipped visits, or the patient not having a wound. In the figures, black vertical lines represent the true parameter value, and bars correspond to 90% credible intervals (CI). We see that the algorithm estimates the parameters correctly, and the CIs cover the true value for all except three parameters, which corresponds to what one would expect from well-calibrated 90% CIs (4*/*44 *≈* 0.09).

**Supplementary Figure 3.**
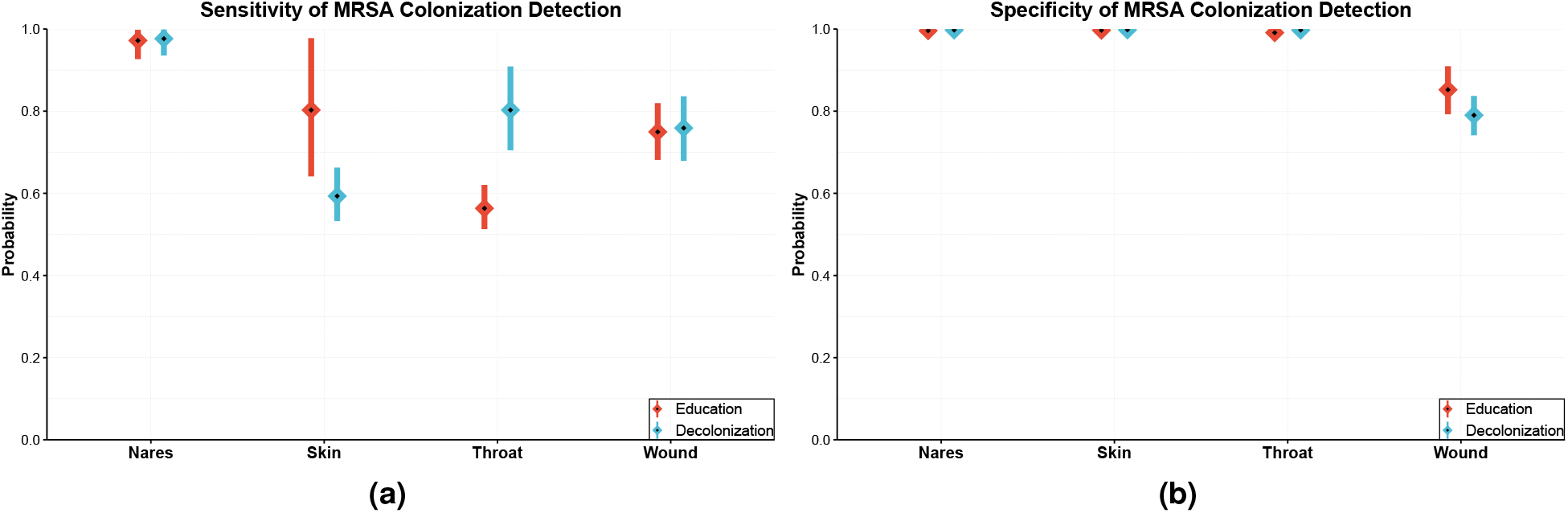
Additional results obtained by the Additive-CHMM: Estimated (a) sensitivity and (b) specificity of MRSA colonization detection by body site. **(a)** Sensitivities (the probability of correctly detected colonized sites) and **(b)** specificities (the probability of correctly detected clear sites) are estimated from the model’s emission probabilities. Calculation of the posterior distributions is explained in the Methods. Means and 90% credible intervals (CI) are represented in the figure by squares and lines, respectively.

**Supplementary Figure 4.**
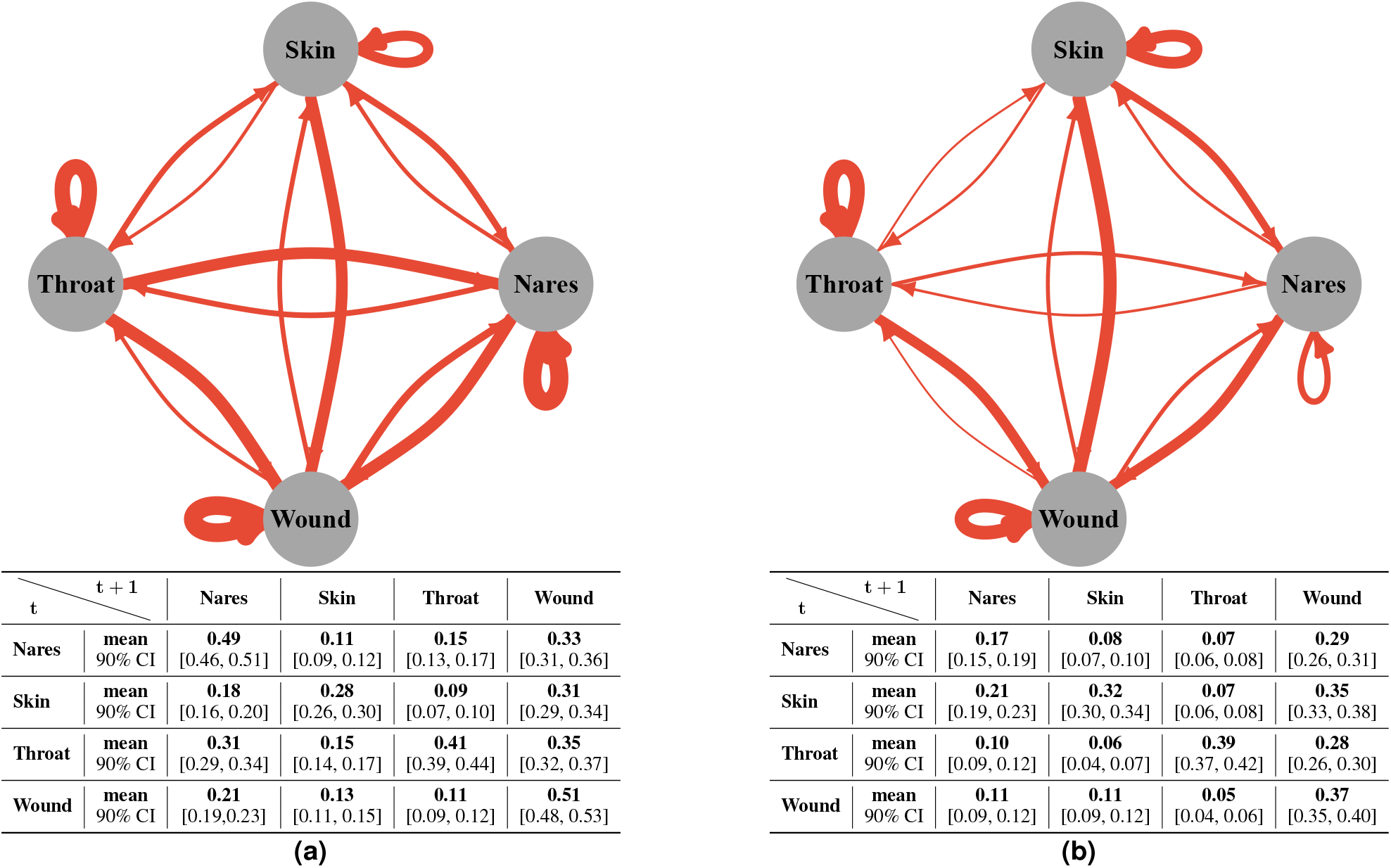
The probability of MRSA transmission among body sites in (a) education and (b) decolonization groups. The edges in the graphs show the probability of an MRSA transmission within a time step (corresponding to 1 month) from the source site to the target conditional on only the source is colonized. They are estimated by simulating a data set with a thousand individuals and considering those patients colonized in the beginning in one site only (i.e., the source of transmission). The edge thickness represents the expected value, and the tables show the means and the respective 90% CIs for all relations.

**Supplementary Figure 5.**
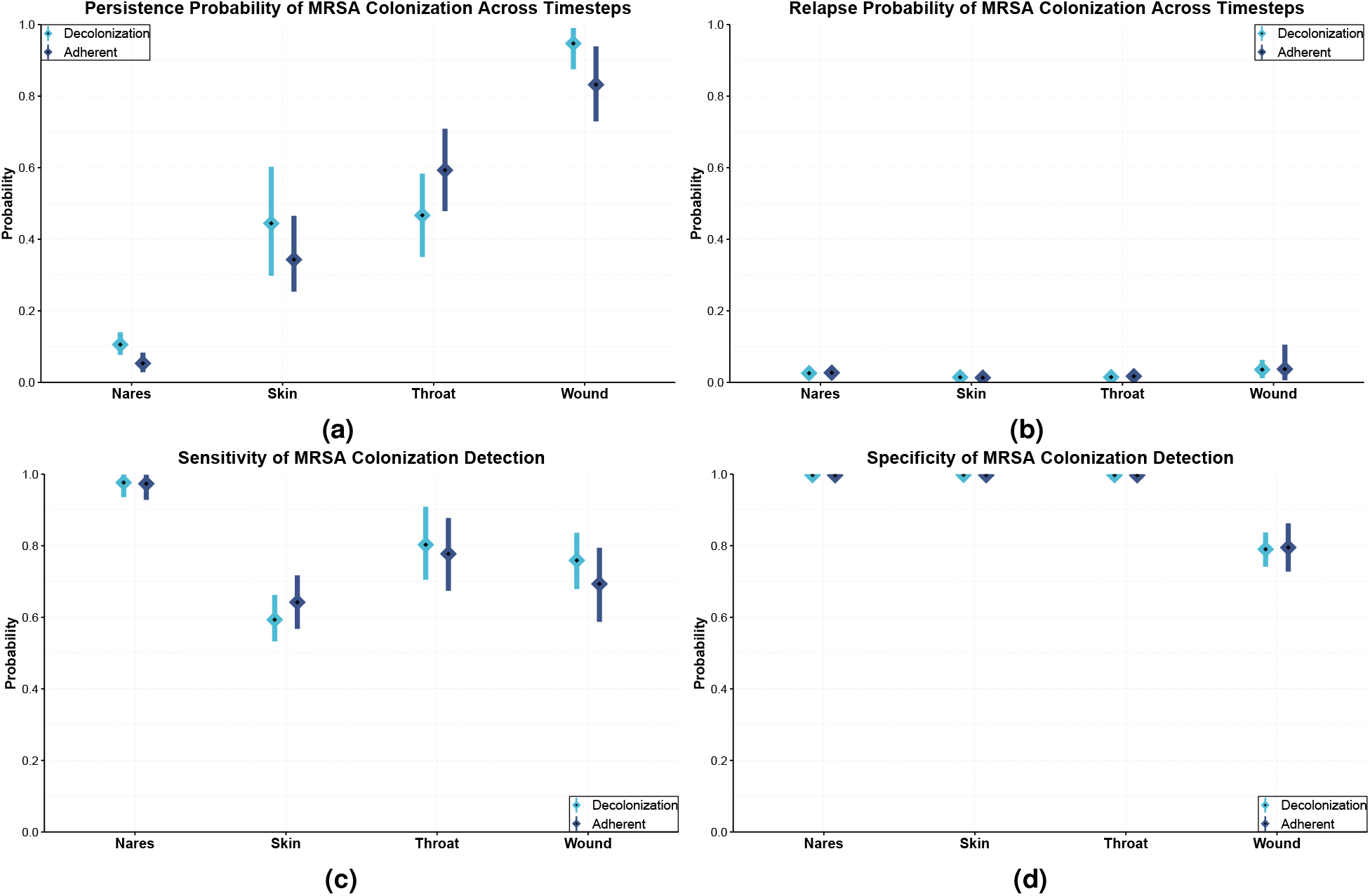
Comparison of fully adherent patients with the full decolonization group regarding Additive-CHMM. The **(a)** persistence probabilities (the probability of the site remaining colonized) and **(b)** relapse probabilities (the probability of the site getting colonized) were estimated from the model’s transition parameters. **(c)** Sensitivities (the probability of correctly detected colonized sites) and **(d)** specificities (the probability of correctly detected clear sites) of MRSA detection were estimated from the model’s emission probabilities. Calculation of the posterior distributions is explained in the Methods. Means and 90% credible intervals (CI) are represented in the figure by squares and lines, respectively.

**Supplementary Figure 6.**
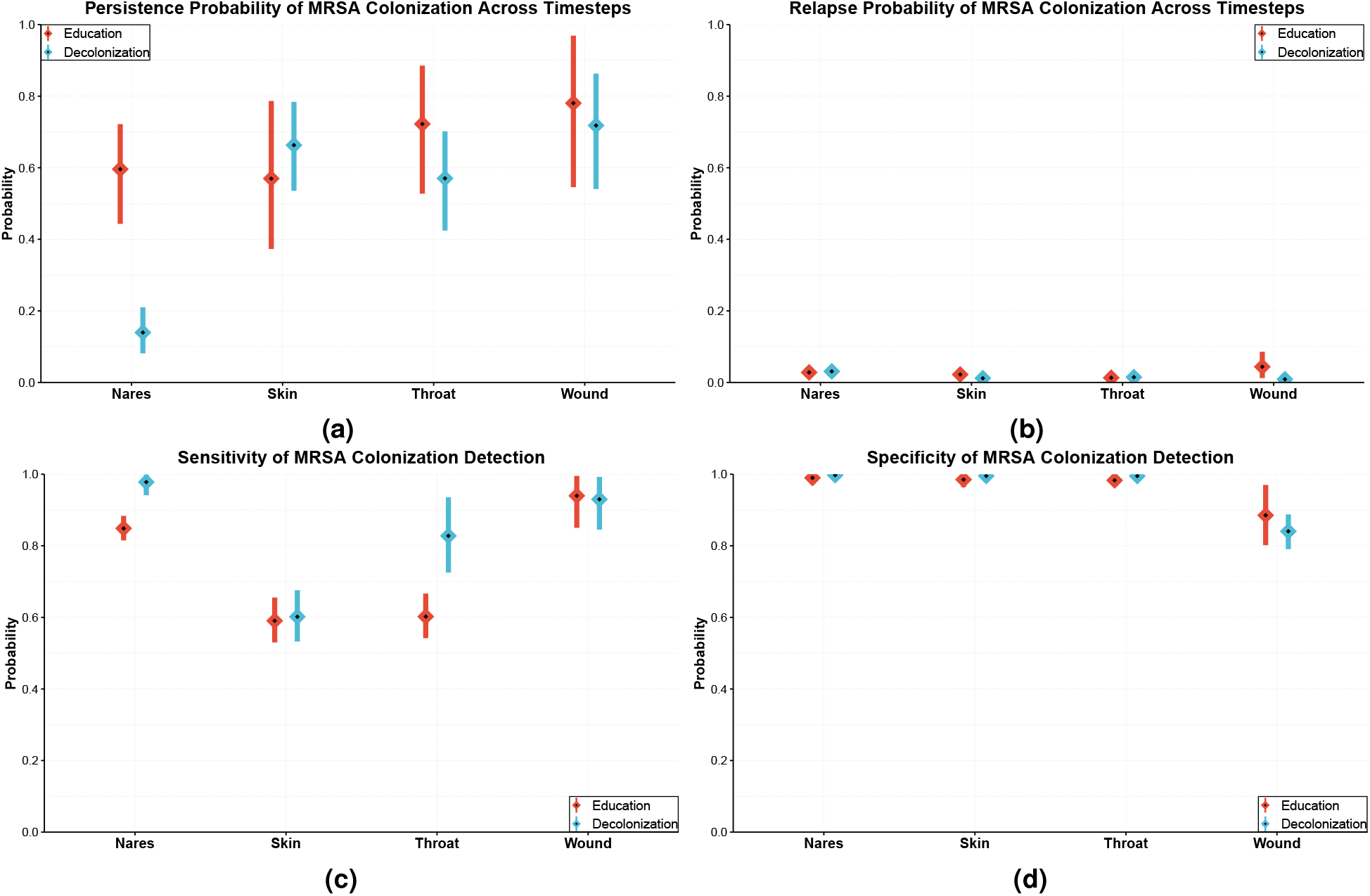
Results of Or-CHMM: Estimated (a) persistence probability and (b) relapse probability of MRSA colonization and (c) sensitivity and (d) specificity of MRSA colonization detection by body site. The **(a)** persistence probabilities (the probability of the site is remaining colonized) and **(b)** relapse probabilities (the probability of the site is getting colonized) are estimated from the model’s transition parameters. **(c)** Sensitivities (the probability of correctly detected colonized sites) and **(d)** specificities (the probability of correctly detected clear sites) are estimated from the model’s emission probabilities. Calculation of the posterior distributions is explained in the Methods. Means and 90% credible intervals (CI) are represented in the figure by squares and lines, respectively.

**Supplementary Figure 7.**
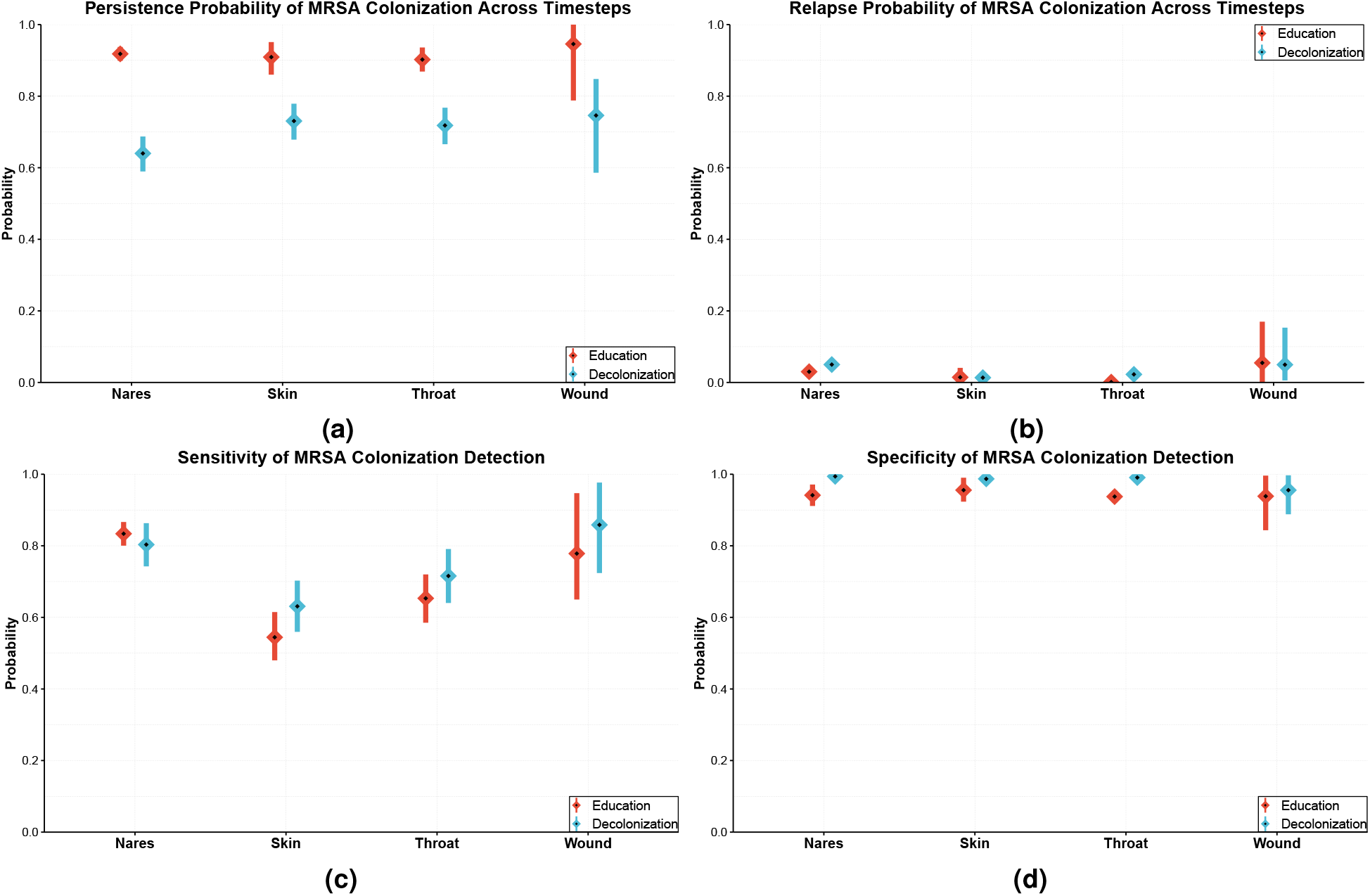
Results of Standard-HMMs: Estimated (a) persistence probability and (b) relapse probability of MRSA colonization, and (c) sensitivity and (d) specificity of MRSA colonization detection by body site. The **(a)** persistence probabilities (the probability of the site is remaining colonized) and **(b)** relapse probabilities (the probability of the site is getting colonized) are estimated from the model’s transition parameters. **(c)** Sensitivities (the probability of correctly detected colonized sites) and **(d)** specificities (the probability of correctly detected clear sites) are estimated from the model’s emission probabilities. Calculation of the posterior distributions is explained in the Methods. Means and 90% credible intervals (CI) are represented in the figure by squares and lines, respectively.

## Footnotes

* Presented priors in the table are not applicable to Standard-HMMs

† Presented model in the main text

## Notes

### Competing Interest Statement

The authors have declared no competing interest.

### Clinical Trial

NCT01209234

### Clinical Protocols

https://doi.org/10.1056/NEJMoa1716771

### Funding Statement

No external funding was received.

### Author Declarations

We used the data from https://doi.org/10.1056/NEJMoa1716771. This trial was approved by the institutional review board of the University of California Irvine. For more details about the data collection, please see the original article https://doi.org/10.1056/NEJMoa1716771

### Summary of Updates

Model has been updated Results are revised and described in more detail Better model convergence analysis has been performed

